# Validation of parent-report questionnaires for large-scale online screening of avoidant/restrictive food intake disorder in children and adolescents

**DOI:** 10.64898/2026.06.25.26356338

**Authors:** Diana Friskson, Felicia Dahlbäck, Laila L.L. Myrberg, Liv Hog, Nadia Micali, Cynthia M. Bulik, Lisa Dinkler

## Abstract

**Objective:** Studies assessing the validity of screening measures for avoidant/restrictive food intake disorder (ARFID) remain scarce. We evaluated the diagnostic performance and validity of an online, parentl⍰reported screening approach for ARFID in a populationl⍰based sample of children.

**Methods:** Participants were drawn from the ARFID Initiative Sweden (ARIES) cohort and included 65 children aged 6–14 years. Parents completed three screening questionnaires—Pica, ARFID, and Rumination Disorder Interview–ARFID Questionnaire (PARDIl⍰ARl⍰Q), Nine-Item ARFID Screen (NIAS), and Parent Eating Disorder Examination Questionnaire (PEDEl⍰Q)—followed by a diagnostic interview (PARDI). Diagnostic performance indices (sensitivity, specificity, positive predictive value [PPV], and negative predictive value [NPV]) were calculated. Convergent validity was assessed via correlations between questionnaire and interview dimensions.

**Results:** The combined screening algorithm demonstrated perfect sensitivity and NPV, indicating accurate detection of all ARFID cases and exclusion of nonl⍰cases. Specificity was high (0.83), and PPVs ranged from 0.91 to 0.95, decreasing to 0.78 under a more conservative operationalization of ARFID Criterion A4 (psychosocial impairment). Diagnostic performance varied across ARFID criteria: PPVs were low for medically anchored Criteria A1–A3 but high for Criterion A4 (psychosocial impairment; PPV□=□0.95). Correlations between screening measures and corresponding interview dimensions were generally moderate to strong, supporting convergent validity. Children meeting threshold ARFID criteria showed significantly greater symptom severity than subthreshold cases at screening.

**Discussion:** These findings support the use of a multil⍰instrument, parentl⍰reported screening approach for ARFID in largel⍰scale pediatric research and highlight the centrality of psychosocial impairment, underscoring the need for standardized operationalization of this criterion.

**Key Points/Summary:**

- A multil⍰instrument, parentl⍰reported online screening approach demonstrated strong diagnostic performance for identifying avoidant/restrictive food intake disorder (ARFID) in a populationl⍰based pediatric sample.
- Diagnostic performance varied across DSM-5 ARFID criteria, with lower accuracy for medically anchored criteria (weight/growth or nutritional impairment) and higher accuracy for psychosocial impairment.
- These findings highlight the utility of this approach for largel⍰scale screening and early identification, particularly in settings where clinical interviews are not feasible.

## Introduction

Avoidant/restrictive food intake disorder (ARFID) is a feeding and eating disorder characterized by avoidance or restriction of food intake—either in quantity or variety—resulting in weight loss or faltering growth, nutritional deficiencies, dependence on nutritional supplements, and/or marked psychosocial impairment. The eating disturbance is typically driven by one or more core dimensions, including heightened sensory sensitivity to food characteristics (e.g., texture, taste, smell), lack of interest in food or eating, or fear of negative consequences such as choking or vomiting (American Psychiatric Association, 2022). ARFID commonly has its onset in early childhood but occurs across the lifespan, affecting both children and adults (D’Adamo et al., 2023; Dinkler et al., 2025; Sader et al., 2023; Sanchez-Cerezo et al., 2023). Given the early onset of ARFID and its potential developmental consequences, reliable and feasible methods for identifying affected children in population-based samples are essential. The present study addresses this need by evaluating the diagnostic accuracy and validity of an online, parent-reported ARFID screening procedure in children aged 6–14 years, using a clinical interview as the reference standard.

Since ARFID was introduced as a diagnostic category in 2013, several assessment tools have been developed. Among the most widely used are the Pica, ARFID, and Rumination Disorder Interview (PARDI; Bryant-Waugh et al., 2019), the PARDI–ARFID Questionnaire (PARDI-AR-Q), developed alongside the PARDI (Bryant-Waugh et al., 2022), and the Nine-Item ARFID Screen (NIAS; Zickgraf & Ellis, 2018). These instruments are available in both self-report and parent-report formats. The PARDI is a semi-structured clinical interview designed to assess the presence and severity of ARFID. Although it demonstrates strong validity (Bryant-Waugh et al., 2019; Cooper-Vince et al., 2022), it requires administration by trained clinicians and is time-intensive, typically taking 45–90 minutes and sometimes up to two hours. This limits its feasibility for large-scale research and epidemiological screening, which rely on well-validated, efficient, and scalable screening tools capable of identifying probable cases and non-cases in a cost-effective and standardized manner. Progress in ARFID research thus depends on the rigorous evaluation of self- and parent-report questionnaires that balance feasibility with diagnostic accuracy in large samples.

Although the PARDI-AR-Q and NIAS are now widely used as screening instruments in clinical and research settings, evidence regarding their criterion validity remains limited. Initial validation work on the PARDI-AR-Q is promising. In a sample of 42 individuals with clinicianl⍰confirmed ARFID and 29 healthy controls, the selfl⍰report version demonstrated a threel⍰factor structure consistent with DSMl⍰5 ARFID dimensions, good internal consistency and convergent validity, and high accuracy in distinguishing cases from nonl⍰cases, with 90% of confirmed ARFID cases screening positive and 93% of controls screening negative (Bryant-Waugh et al., 2022). However, the authors emphasized the need for replication in larger and more diverse samples.

A larger body of research has evaluated the psychometric properties of the NIAS. Studies indicate that adult-, child-, and parent-report versions demonstrate a reliable factor structure corresponding to the three intended subscales across multiple cultural and demographic contexts, as well as expected convergent and divergent validity with measures of related and distinct constructs (Kasak et al., 2024; Medina-Tepal et al., 2023; Ogutlu et al., 2024; Zickgraf et al., 2023). Critically, however, few studies have examined criterion validity of the NIAS against clinical ARFID diagnoses (Billman Miller et al., 2024; Burton Murray et al., 2021; Ortiz et al., 2025), and only one study has done using the parent-reported NIAS (Ortiz et al., 2025).

In a clinically diagnosed sample including individuals with ARFID and other eating disorders, Burton Murray et al. (2021) proposed NIAS subscale cutoff scores to identify ARFID, but these thresholds misclassified approximately half of individuals with other eating disorders as having ARFID. Similar overclassification has been reported by Presseller et al. (2024), who found that 57.0% of individuals with current anorexia nervosa, 30.3% with current bulimia nervosa, and 16.6% with binge-eating disorder screened positive for ARFID on the NIAS. This limitation of misclassification applies to both the NIAS and the PARDI-AR-Q, as neither assesses DSM-5 Criterion C—whether the eating disturbance is better accounted for by another eating disorder—underscoring that they are insufficient as standalone screening tools (Burton Murray et al., 2021; Manwaring et al., 2025; Presseller et al., 2024).

Classification accuracy improves when ARFID screening questionnaires are combined with an additional measure of eating disorder pathology, such as the Eating Disorder Examination Questionnaire (EDE-Q) (Burton Murray et al., 2021). In a recent study, Ortiz et al. (2025) combined positive screening on the NIAS or PARDI-AR-Q with a negative EDE-Q screen and then evaluated criterion validity against the PARDI interview for both self- and parent-report versions of the three questionnaires. They found that the PARDI-AR-Q more accurately identified participants meeting diagnostic criteria for ARFID than the NIAS, and that parent-report versions showed higher accuracy than self-report versions.

The scarcity of studies assessing criterion validity against clinical interview benchmarks highlights the need for further validation of ARFID screening procedures to support accurate case identification. The present study evaluated the diagnostic validity of the online ARFID screening procedure used in the ARFID Initiative Sweden (ARIES). Parent-reported versions of the PARDI-AR-Q, NIAS, and EDE-Q (PEDE-Q) were validated against the parent-reported PARDI interview in children screening positive or negative for ARFID. Criterion validity (accurate ARFID classification) and construct validity (convergence of ARFID dimension scores) were examined.

## Method

### ARFID Initiative Sweden (ARIES)

ARIES is a Swedish cohort study investigating genetic and environmental factors associated with ARFID in children aged 6–14 years (Hog et al., 2025). The cohort comprises 1,671 children who screened positive for ARFID and 661 children who screened negative for ARFID. Participants were recruited via social media, healthcare services, school health services, and advocacy organizations. Enrollment occurred between May 2024 and February 2026. Parents completed online questionnaires assessing eating behavior, health, treatment history, and quality of life. Children provided saliva samples via at-home collection for genetic analyses.

ARIES employed a parent-reported online screening procedure that included the PARDI-AR-Q, NIAS, and PEDE-Q. Screening cases were required to screen positive for ARFID on either the PARDI-AR-Q or the NIAS and screen negative for other eating disorders on the PEDE-Q. Screening controls were required to screen negative and all three questionnaires (**Table 1**).

**Table 1.** Inclusion and exclusion criteria of the ARIES main study (reproduced from Hog et al. 2025).

|  | **Screening cases (ARFID)** | **Screening controls** |
| --- | --- | --- |
| NIAS | Picky Eating score >9 **OR**<br>Low Appetite score >8 **OR**<br>Fear score >9 | Picky Eating score ≤9 **AND**<br>Low Appetite score ≤8 **AND**<br>Fear score ≤9 |
|  | **OR** | **AND** |
| PARDI-AR-Q | Criterion A0 met **AND**<br>Criteria (A1 **OR** A2 **OR** A3 **OR**<br>A4) met | Criteria A0–A4 not met |
|  | **AND** | **AND** |
| EDE-Q | Global score <4 **AND**<br>No vomiting **AND**<br>No laxative use | Global score <4 **AND**<br>No vomiting **AND**<br>No laxative use **AND**<br>Binge eating episodes <5 |
|  | **AND** | **AND** |
| Control questions | No lifetime diagnosis of<br>anorexia nervosa or bulimia<br>nervosa | No lifetime diagnosis of any ED **AND** No<br>first degree relative with a lifetime<br>diagnosis of any ED **AND** No current<br>diagnosis of a psychiatric disorder |

Abbreviations: ARFID, avoidant/restrictive food intake disorder; ED, eating disorder; EDE-Q, Eating Disorder Examination Questionnaire; NIAS, Nine-Item ARFID Screen; PARDI-AR-Q, Pica, ARFID, and Rumination Disorder Interview-ARFID Questionnaire.

*PARDI-AR-Q.* The PARDI-AR-Q is a 32-item questionnaire assessing DSM-5 ARFID Criterion A (A1–A4), the three ARFID dimensions (*Sensory Sensitivity*, *Low interest*, and *Concern* about aversive consequences of eating), and severity of impact (Bryant-Waugh et al., 2022). The parent-report version was used. Dimension scores are calculated as the mean of three Likert scale items (0–6), with higher scores indicating greater severity. Impact severity is assessed as the mean of two Likert scale items (0–6), with higher scores indicating greater severity. Because a one-month time frame is relatively short to capture diagnosed nutritional deficiencies or nutritional supplement prescriptions, the assessment time frame for Criteria A2 and A3 items was extended from one month in the original version to three months in the current study to increase sensitivity. The algorithm used to classify positive/negative ARFID screens on the PARDI-AR-Q is presented in **Table 1**.

*NIAS.* The NIAS is a nine-item questionnaire assessing ARFID dimensions (*Picky Eating*, *Low Appetite*, and *Fear* of aversive consequences) rated on a 0–5 Likert scale (Zickgraf & Ellis, 2018). The parent-report version was used (Ogutlu et al., 2024; Ziółkowska et al., 2022). Subscale scores are calculated as summed item scores, with higher scores indicating greater severity. In the absence of parent-report-specific cutoff scores, we applied those proposed for the self-report version (Burton Murray et al., 2021) (**Table 1**).

*PEDE-Q.* The PEDE-Q is a 29-item parent-report questionnaire assessing non-ARFID eating disorder symptoms and behaviors over the past 28 days (Drury et al., 2023). Items are rated on a 0–6 Likert scale. The PEDE-Q generates four subscale scores (Restraint, Eating Concern, Shape Concern, Weight Concern) and a global score, calculated as mean scores. In addition, the frequency of binge eating and purging behaviors is assessed. The PEDE-Q was used to exclude non-ARFID eating disorders in screening cases and controls (criteria presented in **Table 1**).

### Validation Study

This validation study compared ARIES screening classifications with diagnoses derived from the PARDI clinical interview. From April 2025 onwards, parents who had consented to ARIES within the previous 7 weeks (screening cases, n=111; **Figure 1**) or 11 weeks (screening controls, n=52) were selected to balance age, sex, and screening status and invited by email. Forty-five screening cases (40.5%) and 20 screening controls (38.5%) participated. Participants received two cinema tickets as compensation. Written and oral informed consent were obtained. Interviews were conducted online and recorded. The study was approved by the Swedish Ethical Review Authority (nos. 2023-04638-0; 2025-01356-02).

**Figure 1.**
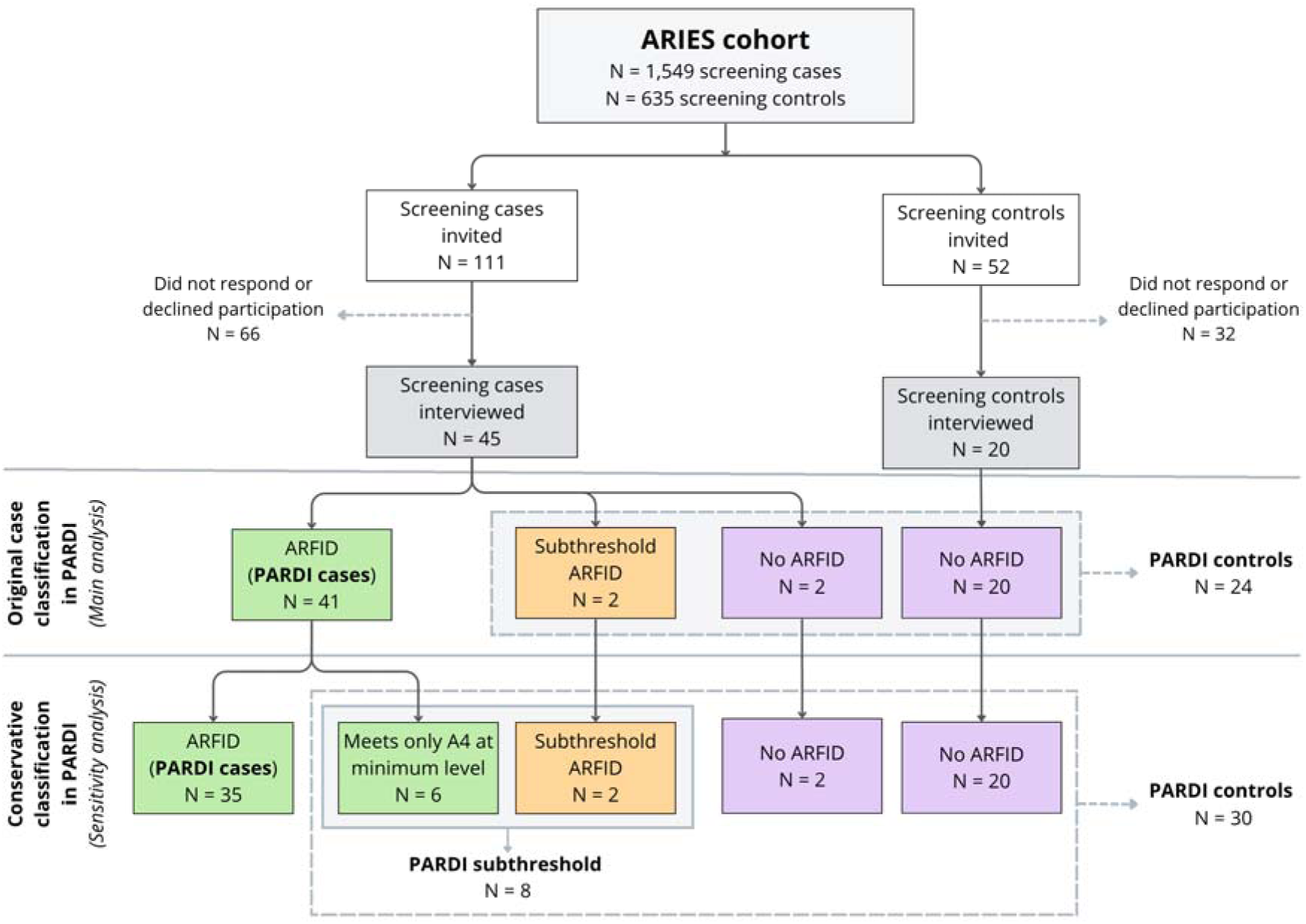
Flowchart of participant recruitment, screening outcomes, and ARFID diagnostic classification. Note: The main analysis used the original classification, in which participants were categorized solely based on PARDI interview outcomes (ARFID dx/Subthreshold ARFID/ No ARFID dx). A sensitivity analysis was performed using an alternative, conservative classification, in which PARDI cases meeting only criterion A4 at the lowest level were reclassified as subthreshold ARFID. In both the original and conservative classifications, the dashed boxes indicate the groups included as PARDI controls. The solid-line box indicates the groups included as subthreshold ARFID for the sensitivity analysis. Abbreviations: ARFID, avoidant/restrictive food intake disorder; ARFID dx, ARFID diagnosis; PARDI, Pica, ARFID and Rumination Disorder Interview; ARIES, ARFID Initiative Sweden

The PARDI is a semi-structured interview assessing all DSM-5 ARFID criteria (A–D) as well as ARFID dimensions and severity (Bryant-Waugh et al., 2019). Question formats are dichotomous or rated on 0–6 Likert scales. Dimension scores (*Sensory sensitivity*, *Low Interest*, *Concern* about aversive consequences of eating) and a *Severity* score are calculated as mean scores. The interview also includes screening for other eating disorders to evaluate Criterion C and assesses medical and psychiatric conditions to evaluate Criterion D (i.e., whether the eating disturbance may be better accounted by another medical or psychiatric condition). Based on PARDI interview outcomes, participants were classified as ARFID (*PARDI case*), no ARFID (*PARDI control*), or subthreshold ARFID (*PARDI subthreshold*). The subthreshold category is not part of the original PARDI but was added in this study for children who did not meet full ARFID criteria yet demonstrated clinically significant, impairing eating problems (applied criteria shown in **Table 2**).

**Table 2.** Criteria applied in the current study to classify PARDI outcomes into *ARFID*, *Subthreshold ARFID*, and *No ARFID*. The subthreshold category was introduced specifically for this study. The original PARDI only differentiates between ARFID and No ARFID.

| DSM-5<br>Criterion | PARDI Question | Required rating |  |  |
| --- | --- | --- | --- | --- |
|  |  | ARFID | Subthreshold<br>ARFID | No ARFID |
| A0 | #29: Do you think your child has an eating or feeding problem that involves avoidance or restriction of food? Has this meant that your child has had difficulty eating enough food overall or has had difficulty eating a wide enough range of foods? | Yes | Yes | No |
| A1 | #34: Over the past 3 months, has your child not put on weight or has he/she been losing weight? Has your child lost weight recently? If so, how much? Have others (e.g., doctors, family) been concerned about this? | ≥4 | 2 or 3 | 0 or 1 |
| A1 | #35: Over the past 3 months has there been concern (e.g., from doctors, family etc.) that your child is not growing taller as he/she should because of their eating habits? | ≥4 | 2 or 3 | 0 or 1 |
| A2 | #36: In the last few months, has your child been identified by a health professional as having any nutritional deficiency due to their eating habits (e.g., low iron, low vitamin B12, low vitamin C)? Who told you this and how did they find out (e.g., blood test)? | Yes | No | No |
| A3 | #37: In the last few months, has any health professional prescribed special supplements for your child (e.g., pills, capsules, powders, or drinks, containing vitamins and or minerals and other micronutrients) specifically to make sure their intake of nutrients is sufficient? | Yes | No | No |
| A3 | #38: Does your child take nutritional supplement drinks (or other high-energy drinks) to help him/her maintain or gain weight? If so, what does he/she take and how much does he/she take each day? | ≥3 | 1 or 2 | 0 |
| A3 | #39: If your child is currently receiving tube feeding, what sort of tube (e.g., nasogastric, PEG [percutaneous endoscopic gastrostomy], or PEG-J [percutaneous endoscopic gastro-jejunostomy])? What is administered via the tube and how much each day? | ≥1 | 0 | 0 |
| A4 | #42: How does your child manage mealtimes? Are they difficult or stressful for them? | ≥4 | 2 or 3 | 0 or 1 |
| A4 | #47: Does your child's eating cause difficulties socially, for example does it make it difficult for him/her to go out with or visit friends, eat at school/college/work, or stay away from home? Has he/she avoided social situations because of eating? | ≥4 | 2 or 3 | 0 or 1 |
| A4 | #48: Does your child's eating cause difficulties in daily functioning at school/college/work? Can you give me examples? | ≥4 | 2 or 3 | 0 or 1 |
| B | #81: The disturbance is better explained by a lack of available food or by an associated culturally sanctioned practice. | No | No | NA |
| C | #82: The respondent has anorexia nervosa or bulimia nervosa, or a related disorder, and there is evidence of a disturbance in the way the individual experiences their body weight or shape. | No | No | NA |
| D | #83: If the respondent has a medical condition, an intellectual disability, other neurodevelopmental disorder, or other mental disorder that explains the eating disturbance, its severity exceeds that usually associated with the condition and requires additional clinical attention? | Yes | Yes | NA |
| Onset | How old was your child when the avoidant or restrictive eating began? | > 1 month | > 1 month | NA |

Abbreviations: ARFID, avoidant/restrictive food intake disorder; NA, Not applicable.

Interviews were conducted via encrypted video calls between April and October 2025 by trained medical and psychology students and one PhD student. The six-hour training included rating a previously recorded interview and completing a supervised interview that was subsequently reviewed. Interviews were discussed in bi-weekly consensus meetings with the study team.

### Statistical Analyses

Analyses were conducted in R (version 4.5.0). Criterion validity was evaluated by comparing ARIES screening classifications with diagnostic outcomes from the PARDI interview. Diagnostic performance was quantified by calculating positive predictive value (PPV; the probability that a child who screened positive truly had ARFID), negative predictive value (NPV; the probability that a child who screened negative was truly free of ARFID), sensitivity (the probability that a child with ARFID was correctly identified as a screening case), specificity (the probability that a child without ARFID was correctly identified as a screening control), and overall accuracy (the proportion of correctly classified cases and controls). These metrics were calculated using the *epiR* package in R (Stevenson & Sergeant, 2025). Height, weight, and body mass index (BMI) are in standard deviations (SD) adjusted by sex and age (Karlberg et al., 2001; Wikland et al., 2002).

Construct validity was assessed by computing Spearman correlation coefficients between continuous dimension scores derived from the PARDI-AR-Q and NIAS and the corresponding dimensions scores obtained from the PARDI interview. Correlations were interpreted as weak (<0.30), moderate (0.30–0.49), or strong (≥0.50), following conventional guidelines (Cohen, 1988). Group differences were examined using Welch’s two-sample *t*-tests for continuous variables, which account for unequal variances between groups, and Pearson’s chi-square tests or Fisher’s exact tests for categorical variables, depending on expected cell counts.

#### Sensitivity Analysis

During consensus meetings, we observed that several children met ARFID diagnostic criteria solely via minimal endorsement of Criterion A4 (psychosocial impairment). Specifically, of the three PARDI items contributing to Criterion A4, only one item was rated 4 on the 0–6 scale—the lowest score exceeding the diagnostic threshold—and none of Criteria A1–A3 (weight/growth or nutritional impairment) were met (**Table 2**). To address potential overclassification of ARFID, these participants were reclassified as having subthreshold ARFID (*PARDI subthreshold*) in a sensitivity analysis (**Figure 1**).

#### Interrater Reliability

Interrater reliability for the PARDI was assessed by double-rating 23 interviews with screening cases (17 parent interviews and 6 child interviews) to ensure consistency and validity across all interviewers when diagnosing ARFID. Cohen’s kappa was calculated for categorical variables and intraclass correlation coefficients (ICC) for continuous variables using the *irr* package in R (Gamer et al., 2019). Interrater reliability was moderate to excellent. Diagnostic agreement for ARFID was 91.3% (κ=0.80, p<0.0001). Agreement was 82.6% for Criterion A1 (κ=0.55, p=0.0085), 100% for Criterion A2 (κ=1.00, p<0.0001), 95.7% for Criterion A3 (κ=0.83, p<0.0001), and 82.6% for Criterion A4 (κ=0.62, p=0.0031). Agreement for the exclusion Criteria B–D was 100%. ICCs indicated excellent reliability for Sensory Sensitivity (ICC=0.89, 95% CI [0.77, 0.95]), Lack of Interest (ICC=0.94, 95% CI [0.87, 0.98]), and Severity (ICC=0.96, 95% CI [0.92, 0.98]), and good reliability for Concern (ICC=0.77, 95% CI [0.54, 0.90]).

## Results

### Sample Characteristics

Sixty-five participants were interviewed with the PARDI: 45 screening cases and 20 screening controls, with a near-equal sex distribution (**Table 3**). Mean age did not differ between screening cases (M=10.5 years) and screening controls (M=10.9 years). The average time from screening to the PARDI interview was approximately seven weeks for screening cases (M=47.2 days) and ten weeks for screening controls (M=67.3 days). In 98.5% of participants, the same caregiver, typically the mother, completed both the screening questionnaires and the PARDI interview.

**Table 3.**
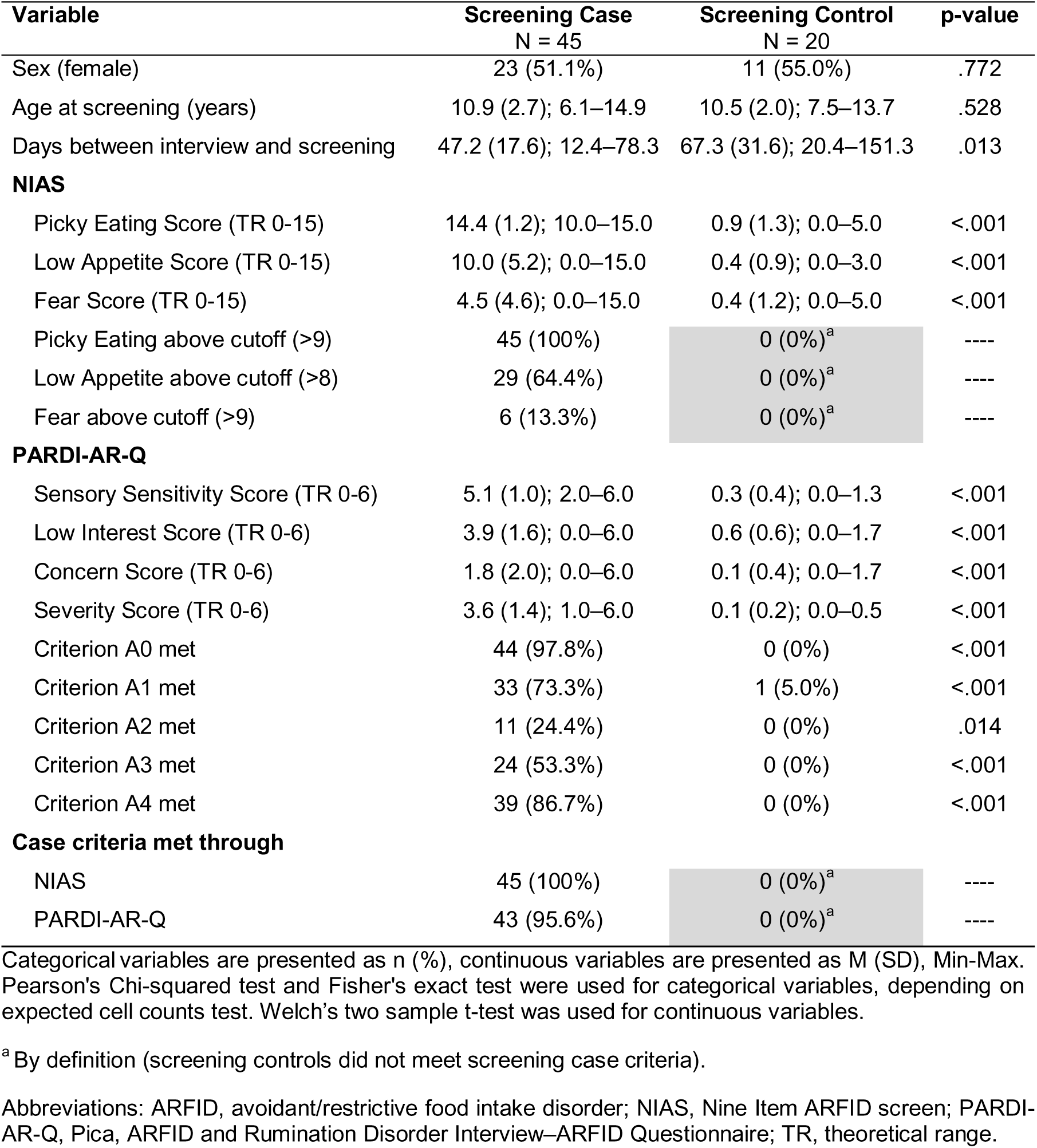
Participant characteristics by screening status.

| Variable | Screening Case<br>N = 45 | Screening Control<br>N = 20 | p-value |
| --- | --- | --- | --- |
| Sex (female) | 23 (51.1%) | 11 (55.0%) | .772 |
| Age at screening (years) | 10.9 (2.7); 6.1–14.9 | 10.5 (2.0); 7.5–13.7 | .528 |
| Days between interview and screening | 47.2 (17.6); 12.4–78.3 | 67.3 (31.6); 20.4–151.3 | .013 |
| **NIAS** |  |  |  |
| Picky Eating Score (TR 0-15) | 14.4 (1.2); 10.0–15.0 | 0.9 (1.3); 0.0–5.0 | <.001 |
| Low Appetite Score (TR 0-15) | 10.0 (5.2); 0.0–15.0 | 0.4 (0.9); 0.0–3.0 | <.001 |
| Fear Score (TR 0-15) | 4.5 (4.6); 0.0–15.0 | 0.4 (1.2); 0.0–5.0 | <.001 |
| Picky Eating above cutoff (>9) | 45 (100%) | 0 (0%)<sup>a</sup> | ---- |
| Low Appetite above cutoff (>8) | 29 (64.4%) | 0 (0%)<sup>a</sup> | ---- |
| Fear above cutoff (>9) | 6 (13.3%) | 0 (0%)<sup>a</sup> | ---- |
| **PARDI-AR-Q** |  |  |  |
| Sensory Sensitivity Score (TR 0-6) | 5.1 (1.0); 2.0–6.0 | 0.3 (0.4); 0.0–1.3 | <.001 |
| Low Interest Score (TR 0-6) | 3.9 (1.6); 0.0–6.0 | 0.6 (0.6); 0.0–1.7 | <.001 |
| Concern Score (TR 0-6) | 1.8 (2.0); 0.0–6.0 | 0.1 (0.4); 0.0–1.7 | <.001 |
| Severity Score (TR 0-6) | 3.6 (1.4); 1.0–6.0 | 0.1 (0.2); 0.0–0.5 | <.001 |
| Criterion A0 met | 44 (97.8%) | 0 (0%) | <.001 |
| Criterion A1 met | 33 (73.3%) | 1 (5.0%) | <.001 |
| Criterion A2 met | 11 (24.4%) | 0 (0%) | .014 |
| Criterion A3 met | 24 (53.3%) | 0 (0%) | <.001 |
| Criterion A4 met | 39 (86.7%) | 0 (0%) | <.001 |
| **Case criteria met through** |  |  |  |
| NIAS | 45 (100%) | 0 (0%)<sup>a</sup> | ---- |
| PARDI-AR-Q | 43 (95.6%) | 0 (0%)<sup>a</sup> | ---- |

Categorical variables are presented as n (%), continuous variables are presented as M (SD), Min-Max. Pearson's Chi-squared test and Fisher's exact test were used for categorical variables, depending on expected cell counts test. Welch's two sample t-test was used for continuous variables.
<sup>a</sup> By definition (screening controls did not meet screening case criteria).
Abbreviations: ARFID, avoidant/restrictive food intake disorder; NIAS, Nine Item ARFID screen; PARDI-AR-Q, Pica, ARFID and Rumination Disorder Interview–ARFID Questionnaire; TR, theoretical range.

All screening cases met NIAS cutoffs on at least one subscale (100% Picky Eating, 64.4% Low Appetite, 13.3% Fear), and all but two (95.6%) screened positive on the PARDI-AR-Q (**Table 3**). On the PARDI-AR-Q, Criterion A4 (psychosocial impairment) was most frequently endorsed (86.7%), followed by Criterion A1 (weight loss/faltering growth; 73.3%), Criterion A3 (dependency on nutritional supplements; 53.5%), and Criterion A2 (nutritional deficiencies; 24.4%).

### Criterion Validity

*ARFID Diagnosis.* Of the 45 screening cases, 41 were confirmed to have ARFID based on the PARDI interview (PARDI cases; PPV=0.91; **Table 4a**). Two of the four non-confirmed screening cases met subthreshold ARFID criteria and were classified as PARDI subthreshold (**Figure 1**). The absence of an ARFID diagnosis was confirmed in all screening controls (PARDI controls; NPV=1.00). Sensitivity, specificity, and overall accuracy of the ARIES screening procedure were 1.00, 0.83, and 0.94 respectively (**Table 4a**). Because all screening cases met NIAS criteria (**Table 3**), these results also reflect the screening performance of NIAS alone (i.e., without the PARDI-AR-Q) when used in combination with the PEDE-Q. We therefore report additional statistics for screening based on the PARDI-AR-Q alone, in combination with the PEDE**-**Q (**Table 4a**). Of the four screening cases ultimately classified as PARDI controls, two screened positive via the NIAS only (i.e., screened negative on the PARDI-AR-Q). Accordingly, the PPV for PARDI-AR-Q-only screening was slightly higher (0.95) than for NIAS-only screening (0.91), although confidence intervals overlapped.

**Table 4.**
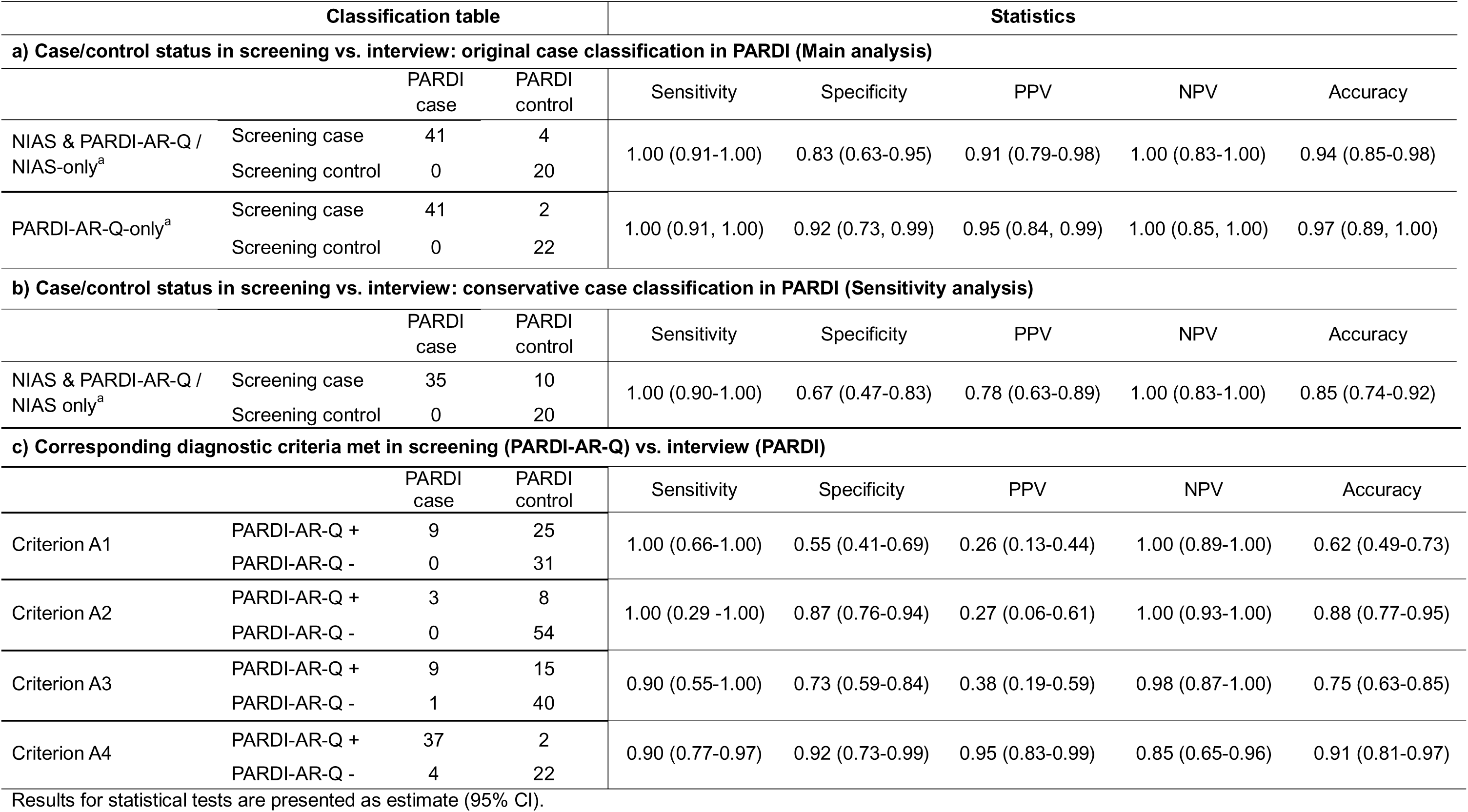
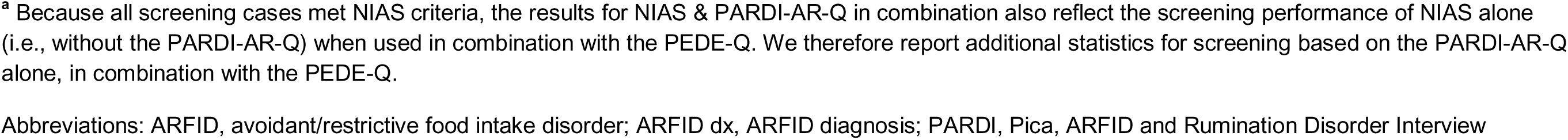
Classification table of agreement between screening and PARDI interview for different case/control definitions and diagnostic criteria.

| Classification table |  |  |  | Statistics |  |  |  |  |
| --- | --- | --- | --- | --- | --- | --- | --- | --- |
| a) Case/control status in screening vs. interview: original case classification in PARDI (Main analysis) |  |  |  |  |  |  |  |  |
|  |  | PARDI case | PARDI control | Sensitivity | Specificity | PPV | NPV | Accuracy |
| NIAS & PARDI-AR-Q / NIAS-only<sup>a</sup> | Screening case | 41 | 4 | 1.00 (0.91-1.00) | 0.83 (0.63-0.95) | 0.91 (0.79-0.98) | 1.00 (0.83-1.00) | 0.94 (0.85-0.98) |
|  | Screening control | 0 | 20 |  |  |  |  |  |
| PARDI-AR-Q-only<sup>a</sup> | Screening case | 41 | 2 | 1.00 (0.91, 1.00) | 0.92 (0.73, 0.99) | 0.95 (0.84, 0.99) | 1.00 (0.85, 1.00) | 0.97 (0.89, 1.00) |
|  | Screening control | 0 | 22 |  |  |  |  |  |
| b) Case/control status in screening vs. interview: conservative case classification in PARDI (Sensitivity analysis) |  |  |  |  |  |  |  |  |
|  |  | PARDI case | PARDI control | Sensitivity | Specificity | PPV | NPV | Accuracy |
| NIAS & PARDI-AR-Q / NIAS only<sup>a</sup> | Screening case | 35 | 10 | 1.00 (0.90-1.00) | 0.67 (0.47-0.83) | 0.78 (0.63-0.89) | 1.00 (0.83-1.00) | 0.85 (0.74-0.92) |
|  | Screening control | 0 | 20 |  |  |  |  |  |
| c) Corresponding diagnostic criteria met in screening (PARDI-AR-Q) vs. interview (PARDI) |  |  |  |  |  |  |  |  |
|  |  | PARDI case | PARDI control | Sensitivity | Specificity | PPV | NPV | Accuracy |
| Criterion A1 | PARDI-AR-Q + | 9 | 25 | 1.00 (0.66-1.00) | 0.55 (0.41-0.69) | 0.26 (0.13-0.44) | 1.00 (0.89-1.00) | 0.62 (0.49-0.73) |
|  | PARDI-AR-Q - | 0 | 31 |  |  |  |  |  |
| Criterion A2 | PARDI-AR-Q + | 3 | 8 | 1.00 (0.29 -1.00) | 0.87 (0.76-0.94) | 0.27 (0.06-0.61) | 1.00 (0.93-1.00) | 0.88 (0.77-0.95) |
|  | PARDI-AR-Q - | 0 | 54 |  |  |  |  |  |
| Criterion A3 | PARDI-AR-Q + | 9 | 15 | 0.90 (0.55-1.00) | 0.73 (0.59-0.84) | 0.38 (0.19-0.59) | 0.98 (0.87-1.00) | 0.75 (0.63-0.85) |
|  | PARDI-AR-Q - | 1 | 40 |  |  |  |  |  |
| Criterion A4 | PARDI-AR-Q + | 37 | 2 | 0.90 (0.77-0.97) | 0.92 (0.73-0.99) | 0.95 (0.83-0.99) | 0.85 (0.65-0.96) | 0.91 (0.81-0.97) |
|  | PARDI-AR-Q - | 4 | 22 |  |  |  |  |  |

Results for statistical tests are presented as estimate (95% CI).
<sup>a</sup> Because all screening cases met NIAS criteria, the results for NIAS & PARDI-AR-Q in combination also reflect the screening performance of NIAS alone (i.e., without the PARDI-AR-Q) when used in combination with the PEDE-Q. We therefore report additional statistics for screening based on the PARDI-AR-Q alone, in combination with the PEDE-Q.
Abbreviations: ARFID, avoidant/restrictive food intake disorder; ARFID dx, ARFID diagnosis; PARDI, Pica, ARFID and Rumination Disorder Interview

*ARFID Criteria A1–A4 (PARDI-AR-Q vs. PARDI)*

Agreement between screening and interview was highest for Criterion A4 (PPV=0.95), whereas considerable discrepancies were observed for Criteria A1–A3 (PPVs=0.26–0.38; **Table 4c**). NPV (0.85–1.00) and sensitivity (0.90–1.00) were high across all criteria. Specificity was high for Criteria A4 (0.92) and A2 (0.87), but lower for A3 (0.73) and A1 (0.55).

### Clinical Characteristics of PARDI Cases and Controls

PARDI cases had significantly lower weight, height, and BMI than PARDI controls (**Table 5**). Only 56.1% of PARDI cases had a BMI in the normal range (-2 to +1 SD), compared with 87.5% of PARDI controls. Among PARDI cases, 24.4% were underweight (< -2 SD; 0% in PARDI controls), and 19.5% were overweight or obese (> +1 SD; 12.5% in PARDI controls).

**Table 5.**
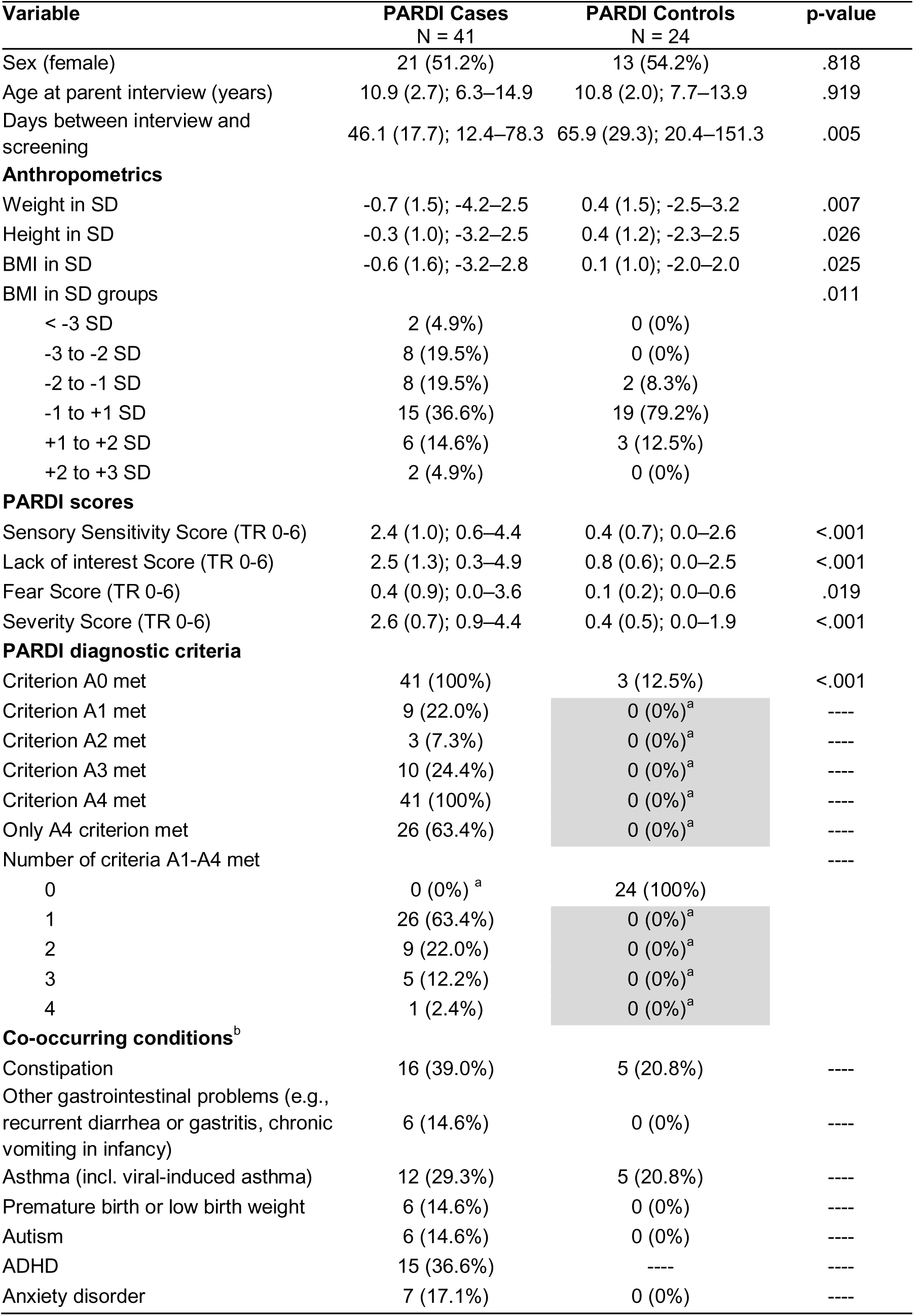
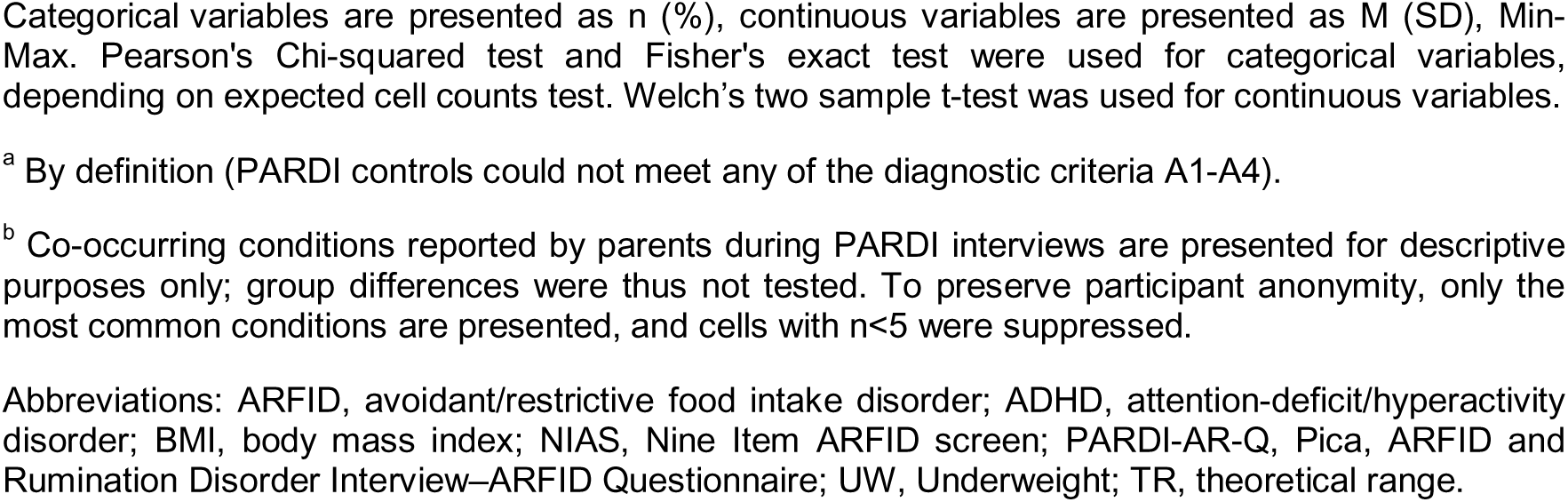
Participant characteristics by PARDI status.

| Variable | PARDI Cases<br>N = 41 | PARDI Controls<br>N = 24 | p-value |
| --- | --- | --- | --- |
| Sex (female) | 21 (51.2%) | 13 (54.2%) | .818 |
| Age at parent interview (years) | 10.9 (2.7); 6.3–14.9 | 10.8 (2.0); 7.7–13.9 | .919 |
| Days between interview and screening | 46.1 (17.7); 12.4–78.3 | 65.9 (29.3); 20.4–151.3 | .005 |
| **Anthropometrics** |  |  |  |
| Weight in SD | -0.7 (1.5); -4.2–2.5 | 0.4 (1.5); -2.5–3.2 | .007 |
| Height in SD | -0.3 (1.0); -3.2–2.5 | 0.4 (1.2); -2.3–2.5 | .026 |
| BMI in SD | -0.6 (1.6); -3.2–2.8 | 0.1 (1.0); -2.0–2.0 | .025 |
| BMI in SD groups |  |  | .011 |
| < -3 SD | 2 (4.9%) | 0 (0%) |  |
| -3 to -2 SD | 8 (19.5%) | 0 (0%) |  |
| -2 to -1 SD | 8 (19.5%) | 2 (8.3%) |  |
| -1 to +1 SD | 15 (36.6%) | 19 (79.2%) |  |
| +1 to +2 SD | 6 (14.6%) | 3 (12.5%) |  |
| +2 to +3 SD | 2 (4.9%) | 0 (0%) |  |
| **PARDI scores** |  |  |  |
| Sensory Sensitivity Score (TR 0-6) | 2.4 (1.0); 0.6–4.4 | 0.4 (0.7); 0.0–2.6 | <.001 |
| Lack of interest Score (TR 0-6) | 2.5 (1.3); 0.3–4.9 | 0.8 (0.6); 0.0–2.5 | <.001 |
| Fear Score (TR 0-6) | 0.4 (0.9); 0.0–3.6 | 0.1 (0.2); 0.0–0.6 | .019 |
| Severity Score (TR 0-6) | 2.6 (0.7); 0.9–4.4 | 0.4 (0.5); 0.0–1.9 | <.001 |
| **PARDI diagnostic criteria** |  |  |  |
| Criterion A0 met | 41 (100%) | 3 (12.5%) | <.001 |
| Criterion A1 met | 9 (22.0%) | 0 (0%)<sup>a</sup> | ---- |
| Criterion A2 met | 3 (7.3%) | 0 (0%)<sup>a</sup> | ---- |
| Criterion A3 met | 10 (24.4%) | 0 (0%)<sup>a</sup> | ---- |
| Criterion A4 met | 41 (100%) | 0 (0%)<sup>a</sup> | ---- |
| Only A4 criterion met | 26 (63.4%) | 0 (0%)<sup>a</sup> | ---- |
| Number of criteria A1-A4 met |  |  | ---- |
| 0 | 0 (0%)<sup>a</sup> | 24 (100%) |  |
| 1 | 26 (63.4%) | 0 (0%)<sup>a</sup> |  |
| 2 | 9 (22.0%) | 0 (0%)<sup>a</sup> |  |
| 3 | 5 (12.2%) | 0 (0%)<sup>a</sup> |  |
| 4 | 1 (2.4%) | 0 (0%)<sup>a</sup> |  |
| **Co-occurring conditions<sup>b</sup>** |  |  |  |
| Constipation | 16 (39.0%) | 5 (20.8%) | ---- |
| Other gastrointestinal problems (e.g., recurrent diarrhea or gastritis, chronic vomiting in infancy) | 6 (14.6%) | 0 (0%) | ---- |
| Asthma (incl. viral-induced asthma) | 12 (29.3%) | 5 (20.8%) | ---- |
| Premature birth or low birth weight | 6 (14.6%) | 0 (0%) | ---- |
| Autism | 6 (14.6%) | 0 (0%) | ---- |
| ADHD | 15 (36.6%) | ---- | ---- |
| Anxiety disorder | 7 (17.1%) | 0 (0%) | ---- |

Categorical variables are presented as n (%), continuous variables are presented as M (SD), Min-Max. Pearson's Chi-squared test and Fisher's exact test were used for categorical variables, depending on expected cell counts test. Welch's two sample t-test was used for continuous variables.
<sup>a</sup> By definition (PARDI controls could not meet any of the diagnostic criteria A1-A4).
<sup>b</sup> Co-occurring conditions reported by parents during PARDI interviews are presented for descriptive purposes only; group differences were thus not tested. To preserve participant anonymity, only the most common conditions are presented, and cells with n<5 were suppressed.
Abbreviations: ARFID, avoidant/restrictive food intake disorder; ADHD, attention-deficit/hyperactivity disorder; BMI, body mass index; NIAS, Nine Item ARFID screen; PARDI-AR-Q, Pica, ARFID and Rumination Disorder Interview–ARFID Questionnaire; UW, Underweight; TR, theoretical range.

Compared with PARDI controls, PARDI cases demonstrated significantly higher scores across all PARDI dimensions and Severity (**Table 5**). Notably, 63.4% of PARDI cases met diagnostic criteria for ARFID exclusively via Criterion A4 (psychosocial impairment). No PARDI cases met diagnostic criteria based solely on Criteria A1, A2, or A3. Criteria B, C and D were met by all screening cases.

PARDI cases also reported a higher number of physical and mental health conditions than PARDI controls, most commonly constipation (39.0%), ADHD (36.6%), asthma (29.3%), and anxiety disorder (17.1%; **Table 5**). None of these conditions precluded an ARFID diagnosis.

### Construct Validity

Correlations between PARDI-AR-Q dimensions and corresponding PARDI dimensions were strong for Sensory Sensitivity (r=0.54) and Low Interest (r=0.67), and moderate for Concern (r=0.47) and Severity (r=0.40; **Figure 2**). Correlations between NIAS subscales and corresponding PARDI dimensions were weak for Picky Eating/Sensory Sensitivity (r=0.19), strong for Low Appetite/Low Interest (r=0.61), and moderate for Fear/Concern (r=0.48). All NIAS subscales showed strong correlations with their corresponding PARDI-AR-Q dimensions (r=0.53–0.85).

**Figure 2.**
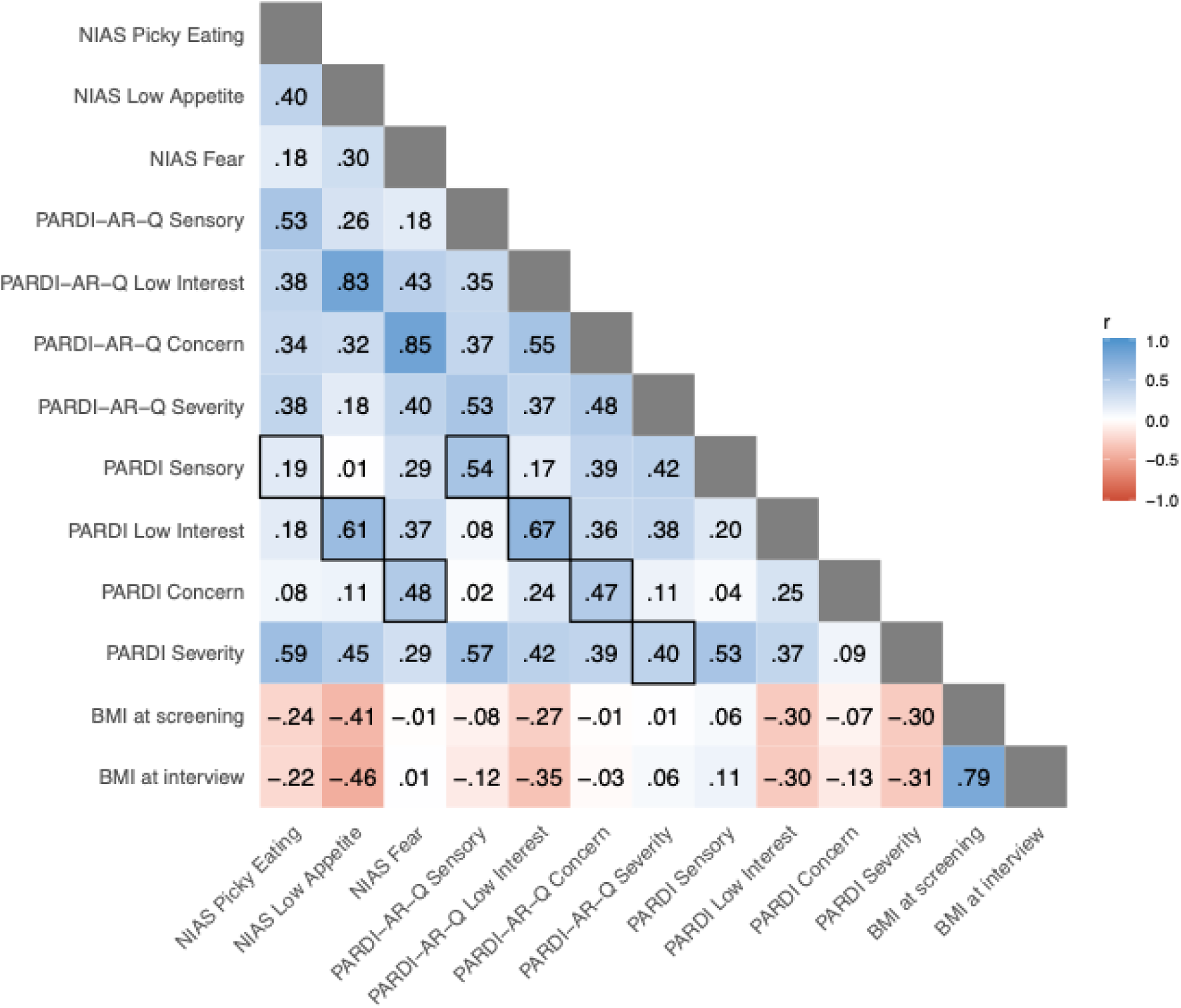
Spearman correlations between parent-reported NIAS profiles (screening), PARDI-AR-Q dimensions (screening) and PARDI dimensions (interview) in screening cases (n=45). Correlation values > 0.5 are bolded. Abbreviations: ARFID, avoidant/restrictive food intake disorder; BMI, body mass index; NIAS, Nine Item ARFID screen; PARDI, Pica, ARFID and Rumination Disorder Interview; PARDI-AR-Q, Pica, ARFID and Rumination Disorder Interview–ARFID Questionnaire

### Sensitivity Analysis

Using the more conservative (i.e., stricter) diagnostic classification (see *Sensitivity Analysis* in the Method section), six participants were reclassified from PARDI case to PARDI subthreshold, reducing the number of confirmed PARDI cases from 41 to 35 and increasing the PARDI subthreshold group to eight children (two originally subthreshold plus six newly reclassified; **Figure 1**). This reclassification reduced the PPV from 0.91 to 0.78. NPV and sensitivity remained at 1.00, whereas specificity decreased from to 0.83 to 0.67, and overall accuracy decreased from 0.94 to 0.85 (**Table 4b**).

Exploratory comparisons showed that the 35 confirmed PARDI cases scored significantly higher than the eight PARDI subthreshold children on the screening measures PARDI-AR-Q Sensory Sensitivity, Concern, and Severity, but not on PARDI-AR-Q Low Interest or any NIAS subscales (**Table 6**). No group differences were observed in age, screening-to-interview interval, or individual PARDI-AR-Q criteria.

**Table 6.** Screening characteristics in confirmed ARFID cases (PARDI Cases) versus PARDI subthreshold participants (conservative classification).

| **Variable** | **PARDI Case**<br>**N = 35** | **PARDI Subthreshold**<br>**N = 8** | **p-value** |
| --- | --- | --- | --- |
| Sex (female) | 19 (54.3%) | 4 (50.0%) | 1.00. |
| Age at screening (years) | 10.9 (2.7); 6.1–14.9 | 10.7 (2.6); 6.5–13.7 | .847 |
| Days between interview and screening | 45.1 (18.9); 12.4–78.3 | 51.0 (6.8); 41.2–60.1 | .155 |
| **NIAS** |  |  |  |
| Picky Eating Score (TR 0-15) | 14.7 (0.7); 12.0–15.0 | 14.1 (1.1); 12.0–15.0 | .173 |
| Appetite Score (TR 0-15) | 10.7 (5.2); 0.0–15.0 | 8.9 (4.6); 2.0–15.0 | .354 |
| Fear Score (TR 0-15) | 5.0 (4.9); 0.0–15.0 | 2.9 (3.2); 0.0–9.0 | .152 |
| **PARDI-AR-Q** |  |  |  |
| Sensory Sensitivity Score (TR 0-6) | 5.4 (0.7); 2.7–6.0 | 4.3 (0.7); 3.0–5.3 | .001 |
| Low Interest Score (TR 0-6) | 4.1 (1.5); 1.0–6.0 | 3.5 (1.4); 0.3–5.0 | .350 |
| Concern Score (TR 0-6) | 2.1 (2.1); 0.0–6.0 | 0.8 (1.1); 0.0–3.0 | .016 |
| Severity Score (TR 0-6) | 3.9 (1.4); 1.0–6.0 | 2.9 (1.1); 1.0–4.0 | .046 |
| Criterion A0 met | 35 (100%) | 8 (100%) | 1.00 |
| Criterion A1 met | 26 (74.3%) | 7 (87.5%) | .656 |
| Criterion A2 met | 9 (25.7%) | 2 (25.0%) | 1.00 |
| Criterion A3 met | 20 (57.1%) | 4 (50.0%) | 1.00 |
| Criterion A4 met | 32 (91.4%) | 6 (75.0%) | .228 |

Subthreshold participants include both the original subthreshold cases and additional participants reclassified based on Criterion A4 at the minimal threshold.
Categorical variables are presented as n (%), continuous variables are presented as M (SD), Min-Max. Fisher's exact test were used for categorical variables. Welch's two sample t-test was used for continuous variables.
Abbreviations: ARFID, avoidant/restrictive food intake disorder; NIAS, Nine Item ARFID screen; PARDI-AR-Q, Pica, ARFID and Rumination Disorder Interview–ARFID Questionnaire; TR, theoretical range.

## Discussion

This study evaluated the diagnostic accuracy and validity of an online screening approach for identifying ARFID in children aged 6–14 years, using a subsample (n=65) from the ARIES cohort (n=2,332). The multi-instrument, parent-reported algorithm combining the PARDI-AR-Q, NIAS, and PEDE-Q demonstrated high diagnostic performance relative to the PARDI interview, supporting its utility in epidemiological research.

The ARIES screening algorithm achieved perfect sensitivity and NPV (i.e., 1.00), indicating that all true ARFID cases were correctly identified and that all who screened negative were truly free of ARFID. High sensitivity is particularly important in population-based screening, where the primary goal is to maximize detection of potential cases and minimize false negatives (Zimmerman, 2024). Specificity was somewhat lower (0.83), reflecting the presence of false positives (children without ARFID screened positive for ARFID); however, this trade-off is generally acceptable in screening contexts, where follow-up diagnostic assessment can clarify case status (Zimmerman, 2024). The high PPVs (0.91 for NIAS+PEDE-Q, 0.95 for PARDI-AR-Q+PEDE-Q, 0.78 for the more conservative case classification) indicate that 78-95% of children who screened positive for ARFID met diagnostic criteria at interview. Together, these findings suggest that combining parent-reported PARDI-AR-Q or NIAS with the PEDE-Q to exclude other eating disorders may provide an effective strategy for large-scale ARFID screening in non-clinical pediatric populations.

Despite strong overall performance, diagnostic accuracy varied substantially across individual ARFID criteria (PARDI-AR-Q vs. PARDI). Criteria A1–A3, reflecting weight/growth or nutritional impairment, demonstrated relatively low PPVs (0.26–0.38), whereas Criterion A4 (psychosocial impairment) showed the strongest diagnostic performance, with a PPV of 0.95. The lower PPVs for Criteria A1–A3 likely reflect differences in how these medically anchored criteria are operationalized in parent-report questionnaires compared with the PARDI interview. For Criterion A1, parents completing screening questionnaires may report minor concerns regarding weight loss or growth faltering that do not meet diagnostic thresholds in the PARDI interview; this applied to 7/25 participants who met Criterion A1 at screening but not in the PARDI. In other instances, parents may primarily be expressing *concern* about potential weight or growth problems rather than reporting objectively verifiable deviations. In contrast, the PARDI requires objectively defined deviations in BMI or growth trajectory (≥0.67 SD decrease) or a markedly low sex- and age-adjusted adult BMI (<17) to meet Criterion A1. Similarly, false positives for Criterion A2 may arise if parents attribute nutritional deficiencies (e.g., Vitamin D deficiency) to eating behavior in the absence of clinical confirmation. Additional discrepancies may reflect misunderstandings of questionnaire instructions, including failure to consider the specified time frame (i.e., past 3 months) or the requirement that supplements be medically prescribed. Notably, during interviews, parents sometimes described clinically relevant symptoms (e.g., dizziness, nausea, fatigue, weakness, or reduced participation in physical activities) despite the absence of a formally diagnosed deficiency, suggesting that parental endorsement may capture functionally meaningful concerns that fall short of formal diagnostic criteria. False screen-positives for Criterion A3 may reflect previous prescriptions of nutritional supplements that were no longer actively used due to child refusal (n=3) or current use of oral nutritional supplement drinks at levels below diagnostic threshold (e.g., 1–2 supplement drinks per day; n=2). In contrast, the PARDI applies a highly stringent criterion, requiring that at least 50% of daily caloric intake be derived from nutritional supplements to meet Criterion A3.

Together, these criterion-level findings underscore the challenges of assessing medically defined ARFID criteria through parent-reported questionnaires alone. In contrast, psychosocial impairment emerged as the most diagnostically robust ARFID criterion in this sample, and it was also the most prevalent one: all children classified as PARDI cases met Criterion A4, and 63% met diagnostic criteria for ARFID based solely on psychosocial impairment. This is consistent with prior research demonstrating high rates of psychosocial impairment among children with ARFID in this age range (Dinkler et al., 2022; Katzman et al., 2021; Watts et al., 2023).

Our findings extend previous work on the criterion validity of ARFID screening measures by evaluating PPV in a populationl⍰based sample that included both screenl⍰positive and screenl⍰negative participants. Bryant-Waugh et al. (2022) demonstrated high concordance between the selfl⍰report PARDIl⍰ARl⍰Q and the PARDI interview in adolescents and adults with clinically established ARFID; however, because diagnostic status was determined prior to questionnaire completion, diagnostic accuracy indices such as sensitivity, specificity, PPV, and NPV could not be estimated, limiting direct comparability with screening studies. More recently, Ortiz et al. (2025) reported PPVs for parentl⍰reported PARDIl⍰ARl⍰Q (0.76) and NIAS (0.64), each evaluated separately against the PARDI interview in a pediatric community sample, making their findings more comparable to the present study. In this context, the substantially higher PPVs observed here for both PARDIl⍰ARl⍰Q (0.95) and NIAS (0.91) warrant consideration of potential methodological explanations.

Differences in PPVs across studies may largely reflect how Criterion A4 (psychosocial impairment) was operationalized and evaluated. In Ortiz et□al., reasons for not meeting ARFID diagnostic criteria at interview were most frequently related to insufficient psychosocial impairment, suggesting that Criterion A4 represented a primary source of diagnostic ambiguity. Consistent with this interpretation, PPV in the present study decreased substantially when a more conservative (i.e., stricter) threshold for Criterion A4 was applied to address potential overidentification related to psychosocial impairment. Specifically, six participants whose scores only marginally exceeded the diagnostic threshold for Criterion A4 in the PARDI were reclassified as having subthreshold ARFID. Under this conservative classification, PPV and specificity decreased to 0.78, whereas sensitivity and NPV remained unchanged, suggesting that the stricter algorithm continued to identify all true ARFID cases while increasing the number of false positives. Importantly, this pattern supports the robustness of the screening approach while also illustrating the sensitivity of diagnostic metrics to how impairment criteria are operationalized, particularly psychosocial impairment, which is inherently subjective and therefore dependent on interpretation. As Harshman et al. (2021) previously demonstrated, prevalence estimates for Criterion A4 can vary dramatically from 18.8% to 86.3% depending on the applied definition. Taken together, these findings suggest that PPV differences across studies are driven primarily by variation in how psychosocial impairment thresholds are defined and applied, underscoring the need for clearer and more standardized assessment guidelines for Criterion A4 in ARFID screening and diagnostic research.

Comparisons between children meeting threshold versus subthreshold ARFID criteria in the PARDI interview revealed clinically meaningful differences. Children meeting threshold ARFID criteria scored significantly higher than subthreshold cases on several PARDI-AR-Q dimensions, indicating differences in symptom severity that were already evident at the screening stage. Prior research suggests that subthreshold ARFID represents a clinically relevant at-risk group. Cooper-Vince et al. (2022) reported elevated ARFID symptom dimensions among subthreshold cases relative to controls, and Kambanis et al. (2025) found that a substantial proportion of individuals with subthreshold presentations later progressed to threshold ARFID. Together with the present findings, this highlights the potential value of screening approaches that identify not only children meeting full diagnostic criteria but also those with subthreshold symptomatology who may benefit from monitoring or early intervention.

Finally, convergent validity analyses further supported the construct validity of the screening measures. As expected, correlations between PARDI-AR-Q dimensions and their corresponding PARDI interview dimensions were moderate to strong (r=0.47–0.67), reflecting their shared conceptual foundation and similar operationalization. Correlations between NIAS subscales and corresponding PARDI dimensions were also moderate to strong for Low Appetite/Low Interest and Fear/Concern (r=0.48–0.61); however, the association between NIAS Picky Eating and PARDI Sensory Sensitivity was weak (r=0.19), consistent with differences in how these constructs are defined and measured. This pattern contrasts with findings by Ortiz et al. (2025), who observed that the parent-reported PARDI-AR-Q Sensory Sensitivity scale showed the weakest convergence with its corresponding PARDI dimension (r=0.31), whereas all other parent-reported correlations of corresponding dimensions were strong (r=0.52–0.67). Notably, Ortiz et al. employed a similar study design and recruitment strategy and included children of comparable ages (7–15 years vs. 6–14 years in the present study). The reasons for these differences remain unclear but may relate to sample-specific differences in symptom distributions or interview scoring practices rather than substantive differences in construct validity across instruments.

### Strengths and Limitations

Key strengths of this study include the evaluation of a prespecified, multil⍰instrument screening algorithm embedded in a reall⍰world, populationl⍰based cohort, which enhances both transparency and ecological validity. Inclusion of both screenl⍰positive and screenl⍰negative participants allowed unbiased estimation of key diagnostic metrics, such as sensitivity, specificity, and NPV, which have rarely been reported in prior validation studies. The study also went beyond categorical diagnosis by assessing construct validity and criterionl⍰level performance, providing nuanced insight into how individual ARFID criteria function in screening contexts. In addition, the larger sample size relative to previous pediatric studies and recruitment from the general population rather than exclusively clinical settings together strengthen the methodological rigor and generalizability of the findings.

Several limitations should be considered. Although larger than previous studies, the sample size remains modest due to the substantial time and personnel demands of the clinical interview, and findings should therefore be interpreted with caution. While recruitment from the general population enhances generalizability, diagnostic performance estimates may be influenced by the distribution of symptom severity, and results may differ in primary care or specialty clinical settings. The study was conducted exclusively in Sweden, and the absence of data on ethnicity and socioeconomic status limits generalizability to more diverse populations; similarly, findings related to psychosocial impairment may not generalize across cultural contexts with different social and eating norms. Results are further limited to children aged 6–14 years and may not extend to other developmental stages. In addition, both screening and diagnostic assessments relied on parent report, raising the possibility of common method variance. Future studies should incorporate multil⍰informant data, including child selfl⍰report or clinicianl⍰observed measures. Voluntary participation may have introduced self-selection bias, with parents of children with more symptoms being more motivated to participate, potentially inflating diagnostic performance estimates. Finally, although interviewers were formally blinded to screening results, screening status may have become apparent during interviews, and interviewer bias cannot be entirely ruled out, potentially leading to an overestimation of diagnostic accuracy.

### Clinical and Research Implications

The present findings support the combined use of parent-reported PARDI-AR-Q, NIAS, and PEDE-Q as a practical and time-efficient screening approach for ARFID in children aged 6–14 years. This multi-instrument strategy appears particularly well suited for large-scale research and for settings in which access to trained clinicians is limited. Future studies should evaluate the diagnostic performance of similar screening approaches in other age groups, validate their use in more diverse cultural and sociodemographic contexts, and further investigate the identification, stability, and clinical significance of subthreshold ARFID presentations. Given the limited existing research on subthreshold ARFID, future work could also examine whether such cases can be reliably identified using empirically derived cutoff scores on parent-reported NIAS or PARDI-AR-Q dimensions.

## Conclusion

This study demonstrates that a multil⍰instrument, parentl⍰reported screening approach can reliably identify probable ARFID and accurately exclude nonl⍰cases in a populationl⍰based sample of children aged 6–14 years. Diagnostic performance remained robust across conservative sensitivity analyses, underscoring the utility of this approach for largel⍰scale screening and early identification efforts where clinical interviews are not feasible. Findings further highlight the central role of psychosocial impairment in pediatric ARFID, as well as the methodological importance of how this criterion is operationalized. Together, the study supports the use of combined screening algorithms for ARFID while underscoring the need for continued validation, particularly with respect to psychosocial impairment thresholds, subthreshold presentations, and application across diverse populations.

## Author Contribution Statement

Friskson: investigation; data curation; visualization; writing – original draft preparation.

Dahlbäck: investigation, data curation; writing – review and editing.

Myrberg: investigation, data curation; writing – review and editing.

Hog: conceptualization; methodology; resources; investigation; data curation; writing – review and editing.

Micali: validation; writing – review and editing.

Bulik: conceptualization; funding acquisition; writing – review and editing.

Dinkler: conceptualization; funding acquisition; methodology; resources; investigation; data curation; formal analysis; visualization; project administration; supervision; writing – review and editing.

## Data Availability

Data and code that support the findings of this study are available from the corresponding author upon reasonable request.

## Ethical Statement

This study was approved by the Swedish Ethical Review Authority (no. 2023-04638-0, 2025-01356-02).

## Acknowledgements

We would like to express our sincere gratitude to the ARFID Initiative Sweden (ARIES) participants, the ARIES Parent Advisory Council, and research nurse Martina Wennberg.

## Disclosure statement about AI use tools

ChatGPT 5.2 was used to write and debug code for data analysis and for proofreading and improving grammar and writing clarity.

## Involvement of persons with lived experiences

The ARFID Initiative Sweden (ARIES) engages a Parent Advisory Council comprising eight parents/guardians of children and adolescents with ARFID to ensure that the research is feasible, relevant, and beneficial to the community. Prior to the main study launch, the Parent Advisory Council consulted with researchers on the selection of questionnaires, completed a beta version of the full study, and provided feedback on all study procedures. The Parent Advisory Council was not involved in the study design, execution, or write-up of the ARIES validation study described here.

